# Optimizing RT-PCR detection of SARS-CoV-2 for developing countries using pool testing

**DOI:** 10.1101/2020.04.15.20067199

**Authors:** Mauricio J. Farfan, Juan P. Torres, Miguel O’Ryan, Mauricio Olivares, Pablo Gallardo, Carolina Salas

## Abstract

The global shortage of reagents and kits for nucleic acid extraction and molecular detection of SARS-CoV-2, requires new cost-effective strategies for the diagnosis of suspected COVID-19 cases, especially in countries that need to increase detection capacity. Pooled nucleic acid testing has been extensively used as a cost-effective strategy for HIV, HepB, HepC and influenza. Also, protocols dispensing of RNA extraction appears as an attractive option for detection of SARS-CoV-2. In this study, pooling nasopharyngeal samples with both automated and manual extraction proved reliable, and thus a potential efficient alternative for the diagnosis of suspected COVID-19 in developing countries.

## Introduction

The pillars for the control of COVID-19 are early detection and quarantine of cases and contacts, and social distancing, especially when detection is suboptimal ^1,2^. Detection is currently based on real time reverse transcriptase-polymerase chain reaction (RT-PCR) in nasopharyngeal samples, which has proven highly specific, and reasonably sensitive during the early symptomatic phase ^3^.

The number of subjects tested for SARS-CoV-2 virus varies significantly by countries, being lowest in developing countries. There are several reasons for this, including lack of a country-based testing strategy, lack of sufficient installed capacity to perform rRT-PCR, and/or lack of reagents to perform a high number of tests due to insufficient supplies of reagents and kits for nucleic acid extraction and molecular detection for SARS-Cov-2 ^4^.

In Chile, as in other middle-high income countries, a country-based strategy aiming to detect as many cases as possible has been implemented. Declared government goals are to increase RT-PCR detection capacity from 1,000 to near 10,000 samples per day for which several strategies have been adopted. The main strategy has been to organize a network of laboratories nationwide, recruiting public and private hospitals and institutions as well as University research laboratories with RT-PCR capacity. These laboratories have worked to develop standardized protocols under government supervision. The main limitation for kick off although has been the availability of reagents. Importantly, the worldwide need for testing allows to envision a shortage of reagents in the short-middle term, which will afflict mostly developing countries.

In this scenario, several research groups are searching for alternative strategies including dispensing of RNA extraction, in-house amplification mixes, and pool testing. Pooled nucleic acid testing has been extensively used as a cost-effective strategy for HIV, HepB, HepC and influenza ^5,6^. We present here results of a pool testing strategy for SARS-Cov-2 including different RNA extraction methods, potentially suitable for developing countries.

## Methods

### Sample pooling

Nasopharyngeal samples in Universal Transport Media (UTM; Copan Diagnostics Inc) from patients COVID-19 positive and negative for SARS-CoV-2 were used. For pooling, 200-µl of 5 nasopharyngeal samples in UTM media were used to create 1-ml pools.

### Nucleic acid extraction

Nucleic acids extraction of 400 µl of the pool of samples was performed using: a) MagNA Pure Compact Nucleic Acid Isolation Kit I (automated extraction) on the MagNA Pure LC instrument or b) High Pure Viral Nucleic Acid Kit (manual extraction), according to the manufacturer’s instructions (Roche). The elution volume was set to 50 µl. We also performed RT-qPCR without extraction, adding 5 µl of the pool samples directly to the RT-PCR reaction.

### SARS-Cov-2 detection

For RT-qPCR detection we used the TaqMan™ 2019-nCoV Assay Kit v1 for the Orf1ab gene, according to the manufacturer’s instructions (ThermoFisher) in a LightCycler® 480 Instrument II (Roche), using 5 µl of nucleic acid extraction or pool samples. A cycle threshold (C_T_) <37 was considered positive.

### Ethical approval

The Molecular Biology laboratory at Calvo Mackenna hospital has been identified by the Health Ministry as a COVID-19 detection site and standardization/improvement of detection methods have been instructed. This work is considered thus for as a public health intervention to improve diagnosis and individual consent nor ethical approval was requested. Nasopharyngeal samples from COVID-19 positive and negative patients were anonymized.

## Results

The Molecular Biology Laboratory at Calvo Mackenna has tested to date 630 samples of which 41 have been positive. For the purpose of this study we selected 23 positive samples with C_T_ ranging from 16.6 to 36.1, and 40 negative samples.

First, we prepared 6 pools of 5 samples subject to automated extraction (Table 1). Pools 2, 4, 5 and 6 included 4 negative and 1 positive samples with C_T_ values of 21.1, 23.8, 26.9 and 31.6, respectively. Pools 1 and 3 contained only negative samples. Amplification of the Orf1ab gene marker was obtained in all pools with a positive sample. The C_T_ values of pools 2, 4, 5 and 6 were 24.3, 27.2, 30.1 and 34, respectively, observing an increase of 2.4 to 3.4 C_T_ units with respect to the C_T_ value of the original sample.

**Table 1.** SARS-CoV2 PCR results obtained from the first six pools of nasopharyngeal samples. Nucleic acids extraction was performed using an automated extraction ^a^

| Sample | C<sub>T</sub> Value<br>SARS CoV-2 | Pool 1 | Pool 2 | Pool 3 | Pool 4 | Pool 5 | Pool 6 |
| --- | --- | --- | --- | --- | --- | --- | --- |
| 1 | Neg | X | X |  |  |  | X |
| 2 | Neg | X | X |  |  |  | X |
| 3 | Neg | X | X |  |  |  | X |
| 4 | Neg | X | X |  |  | X |  |
| 5 | Neg | X |  | X | X |  |  |
| 6 | Neg |  |  | X | X | X |  |
| 7 | Neg |  |  | X | X | X | X |
| 8 | Neg |  |  | X | X | X |  |
| 9 | Neg |  |  | X |  |  |  |
| 10 | 21.1 |  | X |  |  |  |  |
| 11 | 23.8 |  |  |  | X |  |  |
| 12 | 26.9 |  |  |  |  | X |  |
| 13 | 31.6 |  |  |  |  |  | X |
| **SARS-CoV-2 RT-PCR** |  | Neg | Pos | Neg | Pos | Pos | Pos |
| **C<sub>T</sub> value** |  | - | 24.3 | - | 27.2 | 30.1 | 34.0 |
| **ΔC<sub>T</sub>** |  | - | 3.2 | - | 3.4 | 3.2 | 2.4 |

<sup>a</sup> automated extraction was done using MagNA Pure Compact Nucleic Acid Isolation Kit I on the MagNA Pure LC instrument
<sup>b</sup> change in C<sub>T</sub> value compared with C<sub>T</sub> value of the positive sample present in the pool.

For comparison between automated extraction, manual extraction and non-extraction, 5 new pools were prepared (Table 2). Pools 8 to 11 include 4 negative samples and 1 positive sample with C_T_ values of 23.5, 16.8, 26.8 and 35 respectively. Pool 7 included 5 negative samples. SARS-CoV-2 amplification was observed in pools 8, 9, and 10 using automated extraction, manual extraction, or adding the pool sample directly to the PCR mix. Similar C_T_ values were observed using manual or automated extraction, but an increase of ∼5 units was observed by adding 5 µl of non-extracted pool samples to the RT-PCR reaction (Figure 1). There was no amplification signal of SARS-CoV2 in pools 7 (all negative samples) and 11 (4 negative and one positive with a high Ct) for any extraction procedure.

**Table 2.** SARS-CoV-2 PCR results obtained from 5 pools of nasopharyngeal samples. Nucleic acids extraction was performed using an automated ^a^ (A) and manual ^b^ (M) extraction. Adding pool sample (P) directly to PCR reaction was also evaluated ^c^.

| Sample | C<sub>T</sub> Value<br>SARS CoV-2 | Pool<br>7A | Pool<br>8A | Pool<br>9A | Pool<br>10A | Pool<br>11A | Pool<br>7M | Pool<br>8M | Pool<br>9M | Pool<br>10M | Pool<br>11M | Pool<br>7P | Pool<br>8P | Pool<br>9P | Pool<br>10P | Pool<br>11P |
| --- | --- | --- | --- | --- | --- | --- | --- | --- | --- | --- | --- | --- | --- | --- | --- | --- |
| 14 | Neg | X | X | X | X | X | X | X | X | X | X | X | X | X | X | X |
| 15 | Neg | X | X | X | X | X | X | X | X | X | X | X | X | X | X | X |
| 16 | Neg | X | X | X | X | X | X | X | X | X | X | X | X | X | X | X |
| 17 | Neg | X | X | X | X | X | X | X | X | X | X | X | X | X | X | X |
| 18 | Neg | X |  |  |  |  | X |  |  |  |  | X |  |  |  |  |
| 19 | 23,5 |  | X |  |  |  |  | X |  |  |  |  | X |  |  |  |
| 20 | 16,8 |  |  | X |  |  |  |  | X |  |  |  |  | X |  |  |
| 21 | 26,8 |  |  |  | X |  |  |  |  | X |  |  |  |  | X |  |
| 22 | 35 |  |  |  |  | X |  |  |  |  | X |  |  |  |  | X |
| **SARS-CoV-2 RT-PCR** |  | Neg | Pos | Pos | Pos | Neg | Neg | Pos | Pos | Pos | Neg | Neg | Pos | Pos | Pos | Neg |
| **C<sub>T</sub>** |  | - | 25.6 | 18.3 | 29.0 | - | - | 25.2 | 18.5 | 29.0 | - | - | 28.1 | 22.3 | 32.1 | - |
| **ΔC<sub>T</sub>** |  | - | 2.1 | 1.5 | 2.2 | - | - | 1.7 | 1.7 | 2.2 | - | - | 4.6 | 5.5 | 5.3 | - |

<sup>a</sup> automated extraction was done using MagNA Pure Compact Nucleic Acid Isolation Kit I on the MagNA Pure LC instrument (Roche)
<sup>b</sup> manual extraction was done using High Pure Viral Nucleic Acid Kit (Roche)
<sup>c</sup> Five microliters of non-extracted pool samples were added directly to the RT-PCR reaction
<sup>d</sup> change in C<sub>T</sub> value compared with C<sub>T</sub> value of the positive sample present in the pool.

**Figure 1.**
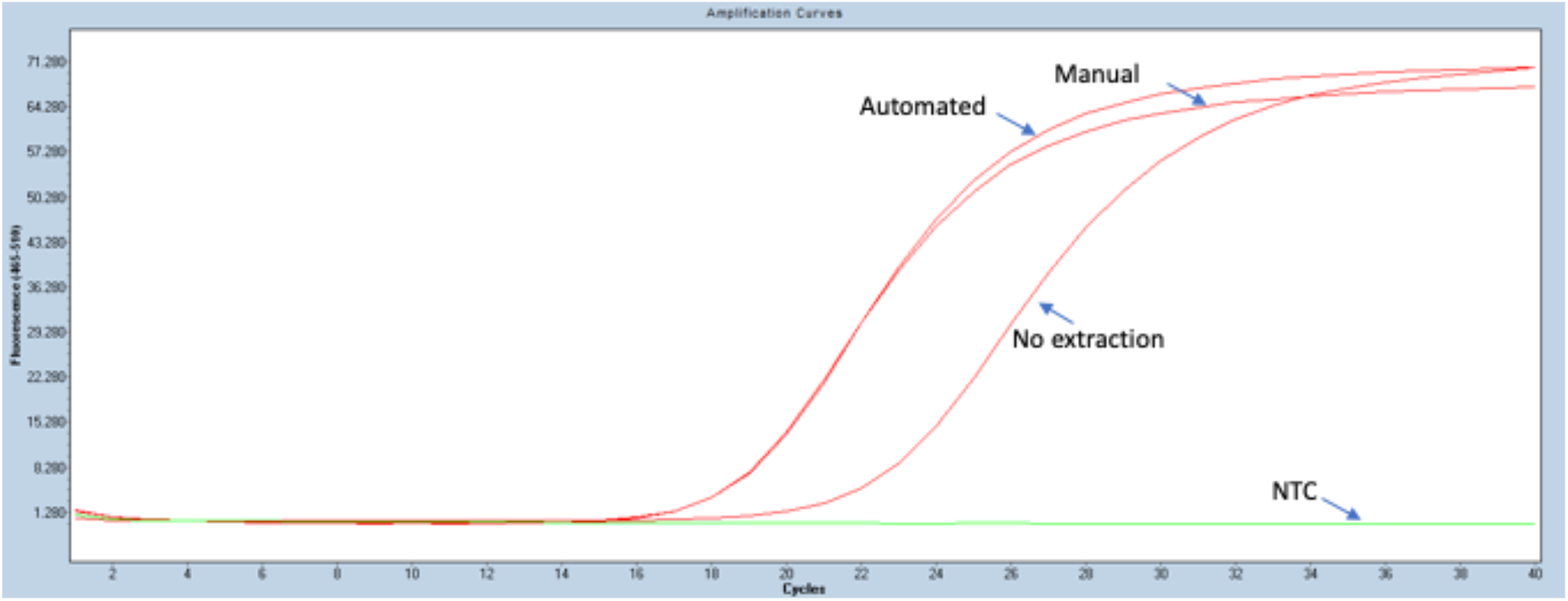
Amplification curves of SARS-CoV-2 obtained for pool 9. SARS-CoV-2 RT-PCR was done using as template nucleic acids purified from automated and manual extraction, or the pool sample (no extraction). NTC, no template control.

To test the efficiency of our previous data, 20 new pools were prepared. Pools 13 to 26 included 4 negative samples and 1 positive sample with C_T_ ranging from 16.6 to 36.1; pools 27 to 31 included 5 negative samples. Extraction of nucleic acid from pools were done by manual extraction. SARS-CoV-2 amplification was observed in pools 13 to 25, observing an increase of C_T_ values from 1 to 4.5. No amplification signal was detected in pool 26, which include a positive sample with a C_T_=36.1. Pools 27 to 31 were all RT-PCR negative (Figure 2).

**Figure 2.**
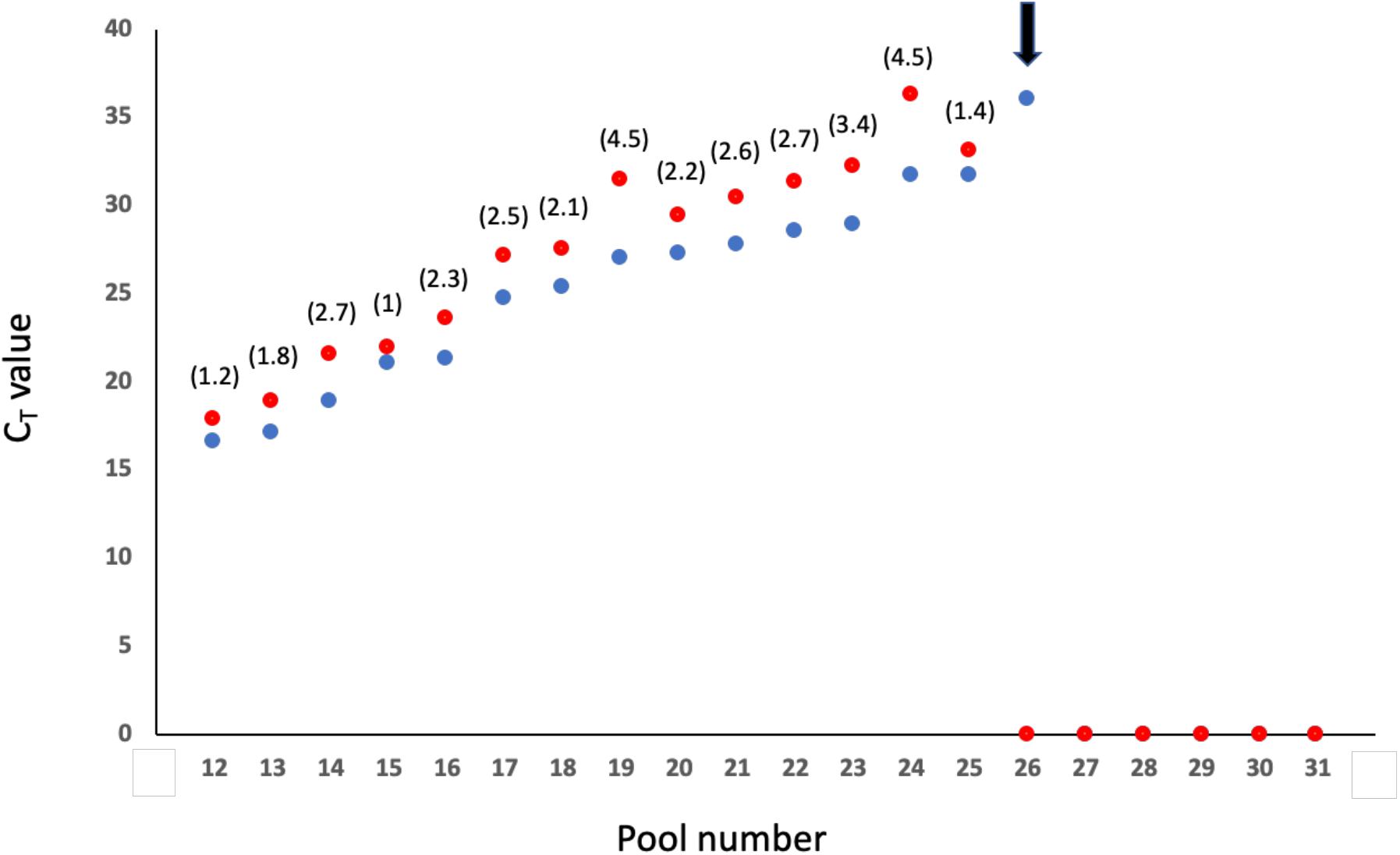
C_T_ values of amplification results of SARS-CoV-2 for pools 12-31. SARS-CoV-2 RT-PCR was done using as template nucleic acids purified from manual extraction and the C_T_ values obtained in the single positive samples (blue dots) and its respective pool (red dots) were graphed. Also, the change in C_T_ value compared with C_T_ value of the single positive sample present in the pool is shown in brackets. A C_T_ value of 0 was assigned to samples with no amplification. Pool 26 is highlighted (black arrow).

## Discussion

Our study demonstrates that pooling of 5 negative and/or 4 negative and one positive SARS-CoV-2 nasopharyngeal samples in the same RT-PCR run can effectively identify all negative samples and detect the positive sample. Furthermore, similar detection results were observed when comparing automated and manual extraction of the sample. Results of the sample without nucleic acid extraction, was unsatisfactory, with a significant increase in C_T_ values, and thus for risk of a false negative result. For extracted samples, C_T_ variations were in the range of 1.0-4,5 units, with less likelihood of a false negative result.

We did not observe significant false negative results. In all the cases in which there was one positive sample, the detection in sample pooling was positive, both in automated and manual extracted samples, except in two cases, where the positive samples have C_T_ values of 35 and 36.1, close to the detection limit of the RT-PCR (C_T_ <37).

Sample pooling has been previously described for the detection of SARS-CoV-2 in developed countries. Hogan et al^7^ studied 292 pools in 2740 nasopharyngeal samples and 148 bronchoalveolar lavage samples, observing a positivity rate of 0.07% in the San Francisco Bay area (CA, USA). This study is complementary to ours as they used samples negative for other viruses, not including samples with known C_T_ values. Another study from Israel, found that a single positive sample could be detected even in pools of extracted nucleic acid of up to 32 samples, with an estimated false negative rate of 10% ^8^.

Multi-sample pools can be a good alternative to increase testing throughput, using less reagents and offering faster results. This is relevant for underdeveloped or developing countries, where resources may be scarce. The possibility of increasing the number of samples for SARS-Cov-2 detection could significantly help countries with reduced resources, to obtain better outcomes for the COVID-19 pandemic. For post-pandemic screening of large populations, sample pooling also will represent an important alternative. Our study has the limitation of having performed only 31 pools on 63 nasopharyngeal samples (40 negatives and 23 positives), however, results were consistent and provide relevant information for the implementation of strategies that might allow optimizing the detection of SARS-CoV-2. We included 5 samples in each pool which seems adequate in our current situation with a near overall 10% positivity rate. In areas with lower positivity rates, especially in future post-pandemic testing, increasing sample numbers in the pool can be considered. Finally, we did not test the inclusion of more than one sample in each pool, however, we would not expect this to modify the observed results.

In conclusion, sample pooling and nucleic acid extraction through automated or manual methods are a reliable and efficient alternative strategy for less developed regions with reduced detection capacity.

## Data Availability

All data referred to in the manuscript is available

